# Ventricular expansion, white matter hyperintensities, and global cognition in Alzheimer’s disease and normal aging

**DOI:** 10.1101/2020.11.30.20240879

**Authors:** Sabrina Adamo, Joel Ramirez, Melissa F. Holmes, Fuqiang Gao, Ljubica Zotovic, Mario Masellis, Sandra E. Black

**Author notes:** ***Correspondence*** Joel Ramirez, PhD, LC Campbell Cognitive Neurology, Sunnybrook Research Institute.

## Abstract

**Background:** The progression of Alzheimer’s Disease (AD) may be tracked by measuring the growth of the ventricular cerebrospinal fluid (vCSF) over time. AD is commonly comorbid with markers of cerebral small vessel disease (SVD), viewed on MRI as white matter hyperintensities (WMH). Larger WMH volumes are correlated with poorer cognitive test scores. Additionally, periventricular WMHs have a proposed relationship to the vCSF.

**Purpose:** This study will examine ventricular expansion and its associations between periventricular/deep WMH and cognition in AD and normal aging.

**Methods:** Baseline and 1-year follow-up data were collected from AD (n=117) and cognitively normal control (NCs; n=49) participants taking part in the Sunnybrook Dementia Study. MRI (1.5T) and scores from both the Mini-Mental State Examination (MMSE) and the Dementia Rating Scale (DRS) were assessed at each time point. Volumetric data was generated using a semi-automated pipeline and each individual’s vCSF and WMHs were transformed to an intermediate space to determine volumetric growth. Regressions were used to determine relationships between vCSF growth measures, SVD burden, and cognition, accounting for demographics and individual interscan intervals.

**Results:** The AD group displayed 14.6% annual ventricular growth as opposed to NC who had only 11.8% annual growth. AD showed significant growth in vCSF (p < 0.001), a trend toward greater pWMH growth (p = 0.06) and no difference in dWMH growth volumes compared to NC. vCSF growth was positively associated with pWMH (β = 0.32, p < 0.001) but not dWMH growth in AD while in NC it was associated with both pWMH (β = 0.48, p < 0.001) and dWMH growth (β = 0.35, p = 0.02). In AD, vCSF growth was associated with the both the MMSE (β = -0.30, p < 0.001) and the DRS (β = -0.31, p < 0.001) in separate models.

**Conclusions:** The findings from this study suggest that in just under 1.5 years, the significantly rapid ventricular expansion observed in AD may be closely related to periventricular small vessel disease. As vCSF growth rates are an important biomarker of AD neurodegeneration that corresponds with cognitive decline, future research should further explore atrophy associated with periventricular vasculopathy.

**Trial Registration:** ClinicalTrials.gov, NCT01800214. Registered on 27 February 2013.

## Background

The progression of neurodegeneration due to Alzheimer’s disease (AD) can be evaluated by ventricular expansion measured from serial magnetic resonance imaging (MRI) [1,2]. As a reliable indicator of brain atrophy and disease progression, ventricular expansion may be a useful biomarker for clinical trials.

In a mixed dementia population, participants with larger ventricles showed signs of lower cognitive reserve [3]. Specifically in AD, ventricular atrophy has been linked with cognitive impairment and future cognitive decline [4]. Moreover ventricular expansion rate has been shown to differ between normal aging, MCI, and AD demonstrating its utility as a marker for progression along the disease spectrum [5,6].

Additionally, correlations with MRI-based markers of cerebral small vessel disease (SVD), measured by white matter hyperintensity (WMH) burden, may also be helpful predictors of future brain atrophy and disease progression [7–9]. The National Institute on Aging-Alzheimer’s Association (NIA-AA) core clinical criteria [10] for the diagnosis of probable/possible AD allows for minimal burden of WMH, however in a real-world community-based and memory clinic setting, SVD is often seen comorbidly with AD [11].

Both AD and SVD pathologies are highly concurrent [8,12] and greater WMH load has also been linked with late onset AD [13]. Previous findings suggest periventricular (pWMH) moreso than deep WMH (dWMH), may be more closely related to ventricular expansion due to its hypothesized role as the cerebrospinal fluid (CSF) circulation and waste clearance in the brain [14].

In addition to vCSF growth, WMH burden has also been linked with cognitive decline in AD [15]. Furthermore, patients with concomitant AD and vascular pathologies such as atherosclerosis (large blood vessels) and arteriolosclerosis (small blood vessels) have been shown to have lower global cognitive test scores [16].

This study will examine the relationship between changes in vCSF/WMH and cognition within the setting of AD and normal aging, determining its validity as a biomarker for tracking disease progression. The two hypotheses for this study are: 1) vCSF growth will be correlated with pWMH but not dWMH; and 2) baseline cognitive test scores will predict vCSF growth in AD but not normal aging.

## Methods

### Participants

Study participants were recruited as part of the Sunnybrook Dementia Study (SDS; ClinicalTrials.gov NCT 01800214) conducted at the Linda C. Campbell Cognitive Neurology Research Unit, Hurvitz Brain Sciences Program at Sunnybrook Research Institute, University of Toronto, Canada. Patients with Alzheimer’s disease (AD: n=117) as well as cognitively normal controls (NC: n=49) with longitudinal neuroimaging and neuropsychological testing were included in the study. All AD patients met the National Institute on Aging - Alzheimer’s Association criteria [10] for probable/possible AD. Participants with a history of head trauma, substance abuse, overt stroke, tumours, or other mental illness unrelated to dementia were excluded, however, the presence of white matter hyperintensities on MRI were not exclusionary for both AD and NC participants. NCs were required to meet strict guidelines based on neuropsychological testing to ensure they were within normal cognitive limits for their age and education levels. Among other neuropsychological assessments, all participants were tested on the Mini-Mental State Examination (MMSE) and the Dementia Rating Scale (DRS), both of which assess global cognitive functioning. Only participants with an inter-scan interval (ISI) within 1-3 years were included in the study. Research was ethically conducted and approved by the Sunnybrook Research Ethics Board.

### MRI procedures

Standardized brain imaging protocol was acquired on a 1.5T GE Signa scanner (Milwaukee, WI, USA) with a mean ISI of 1.3 years (Table 1). Detailed information regarding MRI acquisition protocol are previously published [17].

**Table 1.** **Demographics and raw baseline volumetric data**.

|  | AD<br>(n = 117) | NC<br>(n = 49) | P | Cohen's d<br>effect size |
| --- | --- | --- | --- | --- |
| **Baseline Demographics** |  |  |  |  |
| Age, years | 71.7 (8.5) | 69.6 (7.6) | 0.14 | 0.26 |
| Sex, n (M / F) | 48 / 69 | 21 / 28 | 0.86† |  |
| Education, years | 13.5 (3.4) | 15.8 (2.7) | 0.00 | 0.75 |
| Inter-scan Interval, years | 1.3 (0.4) | 1.4 (0.4) | 0.75 | 0.25 |
| MMSE/30<sup>a</sup> | 23.1 (2.8) | 28.8 (1.3) | 0.00 | 2.61 |
| DRS/144<sup>a</sup> | 118.7 (9.7) | 140.9 (2.1) | 0.00 | 3.16 |
| **Baseline Volumetrics** |  |  |  |  |
| ST-TIV, cc | 1189 (139.7) | 1224.8 (116) | 0.12 | 0.28 |
| BPF, % | 73.2 (4.3) | 78.7 (4.1) | 0.00 | 1.31 |
| vCSF, cc | 49 (21.5) | 34.8 (17.8) | 0.00 | 0.72 |
| sCSF, cc | 271.6 (64.4) | 227.4 (52.7) | 0.00 | 0.75 |
| NAGM, cc | 503.2 (54) | 556.1 (41.6) | 0.00 | 1.92 |
| NAWM, cc | 357.4 (50.5) | 402.3 (51.3) | 0.00 | 0.88 |
| pWMH, mm<sup>3</sup> | 6602.2 (8076.5) | 3754.7 (5522.9) | 0.03 | 0.41 |
| dWMH, mm<sup>3</sup> | 1015.7 (1201.1) | 571.1 (617.2) | 0.02 | 0.47 |
| Right hippocampus<sup>c</sup>, mm<sup>3</sup> | 2642.3 (411.2) | 3286.6 (311.8) | 0.00 | 0.81 |
| Left hippocampus<sup>c</sup>, mm<sup>3</sup> | 2485.2 (470.8) | 3182.5 (323.5) | 0.00 | 0.88 |

All values are presented as mean (SD) unless otherwise indicated. All volumetric data is presented either in cubic centimeters (cc) or cubic millimeters (mm<sup>3</sup>) as indicated in the table. MMSE = Mini-Mental State Examination, DRS = Dementia Rating Scale, ST-TIV = Supratentorial Total Intracranial Volume, BPF = Brain Parenchymal Fraction, vCSF = ventricular Cerebrospinal Fluid, sCSF = sulcal Cerebrospinal Fluid, NAGM = Normal Appearing Grey Matter, NAWM = Normal Appearing White Matter, pWMH = periventricular White matter Hyperintensities, dWMH = deep White Matter Hyperintensities. † = Chi squared, <sup>a</sup> = based on n=112 (AD), n=49 (NC), <sup>b</sup> = based on n=111 (AD), n=49 (NC), <sup>c</sup> = based on n=113 (AD), n=48 (NC)

The full methods used to extract volumetric imaging data on an individual basis have been previously described [18–20]. In summary, the Supratentorial Total Intracranial Volume (ST-TIV) was segmented into grey matter (GM), white matter (WM), sulcal (sCSF) and ventricular cerebrospinal fluid (vCSF) using an automated histogram-based segmentation tool [21]. White matter hyperintensities (WMH) were then classified using a tri-feature (T1/PD/T2) segmentation tool and automatically sub-classified WMH into either deep (dWMH), periventricular (pWMH), or lacunes [19].

The dynamic progression method is previously published [17]. In brief, participant baseline and follow-up vCSF and WMH segmentations were registered to an intermediate space then masked using the ‘fslmaths’ command [22] to generate spatial difference and overlap maps. Spatial difference maps included the growth (“grow”, where volume existed in follow-up but not in baseline), shrinkage (“shrink”, where volume existed in baseline but not in follow-up), and spatial overlap maps (“stable”, common in both baseline and follow-up) (**Figure 1**).

**Figure 1.**
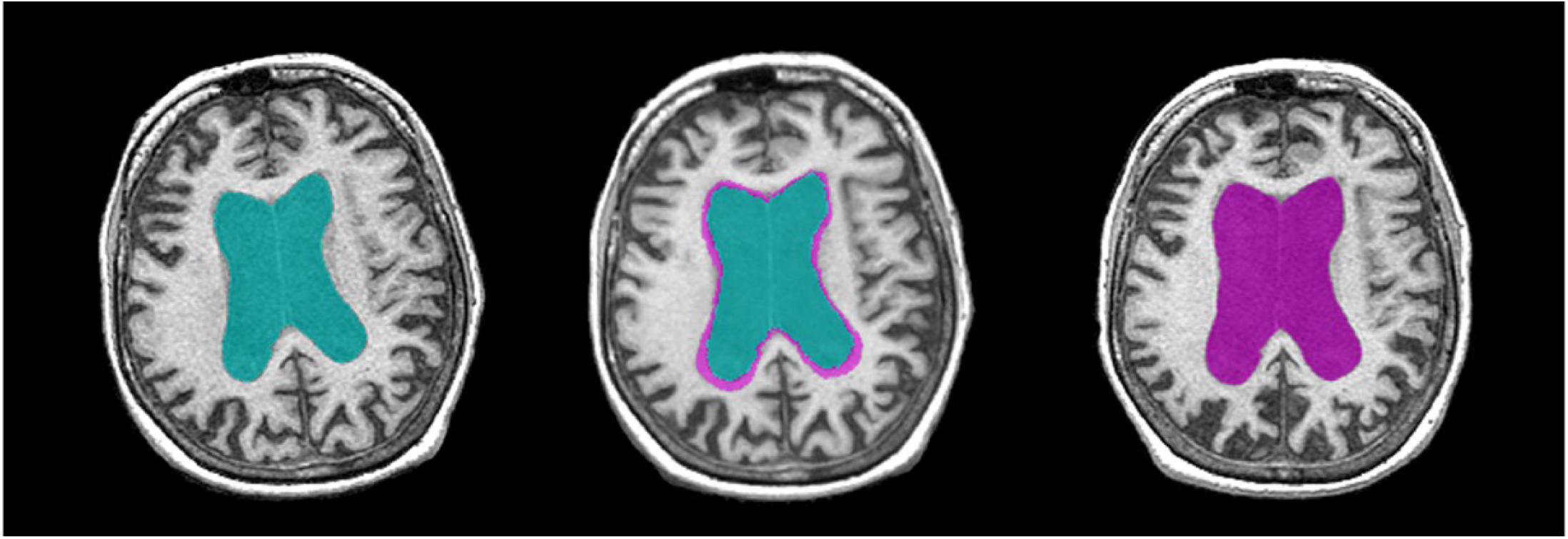
T1 image of the same patient at baseline (left), follow up (right), and an intermediate space (center). Baseline ventricles are shown in green and follow up in purple.

### Statistical Analysis

IBM SPSS (version 24) was used to run all statistical analyses. The mean raw volumes were displayed for transparency, however statistical analyses were run on normalized variables where applicable. One-way Analysis of Variance (ANOVA) was used to test for between-group differences. A Pearson’s chi-squared test was used to examine between-group sex differences. Effect sizes were determined using Cohen’s d. Linear regressions were used to determine the relationships between ventricular growth, cognition and SVD burden. Two models were tested using either the MMSE (Model 1) or DRS (Model 2) to assess for cognition.

## Results

Demographic information is presented in **Table 1**. There was no significant difference between AD and NC in age, sex, interscan interval (ISI), or Supratentorial Total Intracranial Volume (ST-TIV). The AD cohort had significantly lower cognitive test scores on both the MMSE (p < 0.001) and DRS (p < 0.001) than the NC cohort. AD also had a significantly smaller Brain Parenchymal Fraction (BPF; p < 0.001), as well as lower volumes for Normal Appearing Grey Matter (NAGM; p < 0.001), Normal Appearing White Matter (NAWM; p < 0.001), and both right (p < 0.001) and left (p < 0.001) hippocampal volumes. AD had greater volumes of vCSF (p < 0.001), sCSF (p < 0.001), pWMH (p = 0.03), and dWMH (p = 0.02).

### Main finding

Dynamic progression analyses (**Table 2**) revealed that in just under 1.5 years, AD patients exhibited a greater increase in vCSF growth (p < 0.001, **Figure 2c**) and a trend toward greater pWMH growth (p = 0.06) while controlling for age, sex, education, and ISI (**Figure 2f**). There was no significant growth of dWMH volumes (**Figure 2i**). One AD participant was removed from this analysis as a pWMH outlier, bringing the sample to 116. Similar results were also obtained when running the analyses after removing the outlier from the AD group (**Figure 2f**).

**Table 2.** **Ventricular and White Matter Hyperintensity Progression**

|  | AD<br>(n = 116) | NC<br>(n = 49) | P | Cohen's d<br>effect size |
| --- | --- | --- | --- | --- |
| **Growth Volumes (cc)** |  |  |  |  |
| vCSF | 8.9 (4.9) | 4.7 (1.5) | 0.00 | 1.16 |
| pWMH | 3.1 (3.6) | 1.6 (1.9) | 0.06 | 0.52 |
| dWMH | 0.6 (0.6) | 0.4 (0.3) | 0.84 | 0.42 |

All growth volumes are presented as mean (SD) in cubic centimeters (cc). pWMH = periventricular White matter Hyperintensities, dWMH = deep White Matter Hyperintensities.

**Figure 2.**
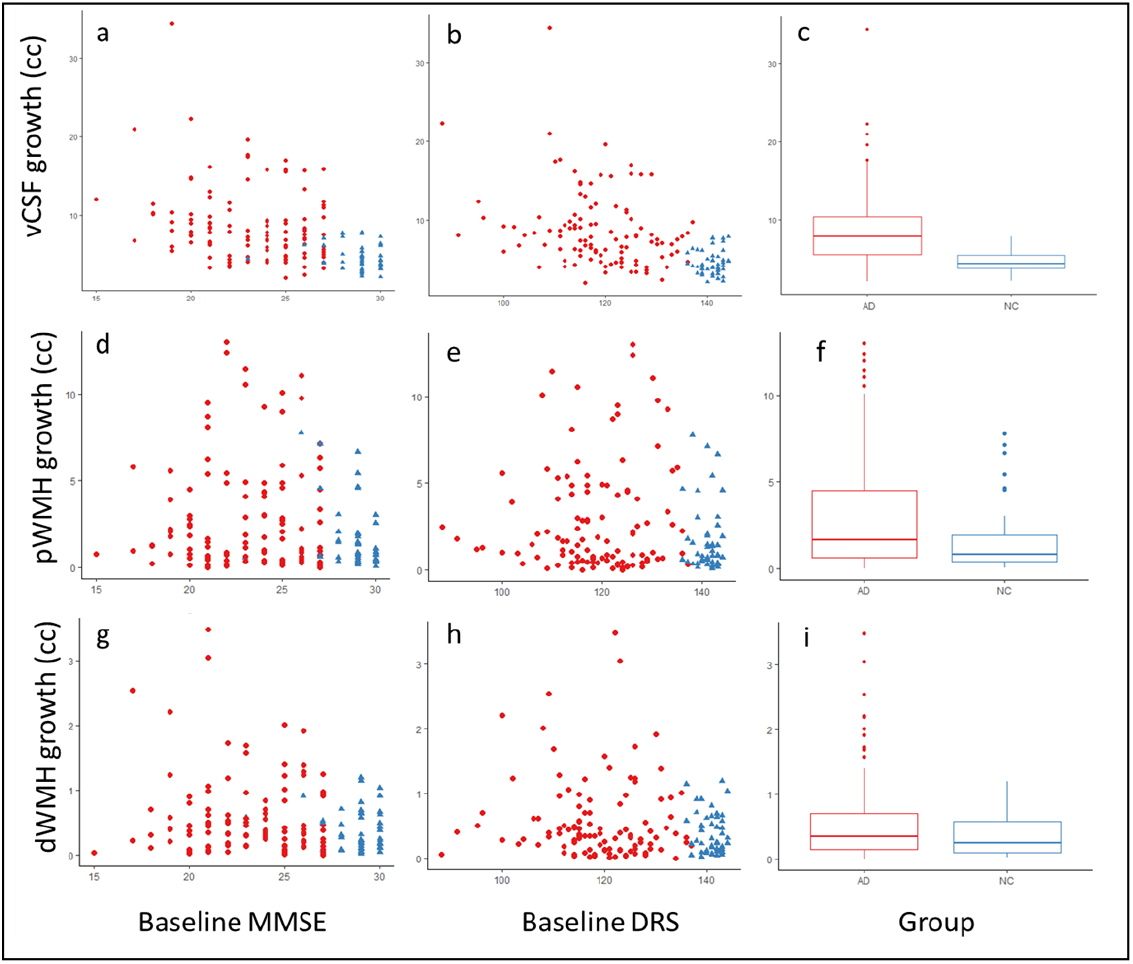
Ventricular growth (top), periventricular (middle), and deep (bottom) White Matter hyperintensities plotted against baseline MMSE and DRS scores. Box plots showing the distribution of each respective growth volumes between diagnostic groupings. The AD cohort is in red and NCs are blue.

### Is vCSF growth associated with SVD burden?

In AD patients, vCSF growth was positively associated with pWMH (β = 0.32, p < 0.001) but not with dWMH growth while correcting for Years of Education (YOE), ISI, and MMSE. ISI was associated with dWMH (β = 0.23, p = 0.03) but not pWMH. NCs, however showed significant associations between vCSF growth and both pWMH (β = 0.48, p < 0.001) and dWMH growth (β = 0.35, p = 0.02). YOE was negatively associated with pWMH in the NC cohort (β = -0.37, p = 0.01).

### Do baseline cognitive test scores predict ventricular growth?

Model 1: In the AD group, vCSF growth showed a negative association with baseline MMSE (β = -0.30, p < 0.001, **Figure 2a**), with ISI (β = 0.40, p < 0.0001) and YOE (β = 0.24, p < 0.01) showing significant effects as well. For NCs, vCSF growth was not associated with MMSE, although there were significant associations with age (β = 0.35, p = 0.02) and sex (β = -0.33, p = 0.01) (**Table 3**).

**Table 3.** **Summary of regressions**

|  |  | dWMH growth |  | pWMH growth |  | vCSF growth - Model 1 |  | vCSF growth - Model 2 |  |
| --- | --- | --- | --- | --- | --- | --- | --- | --- | --- |
|  |  | β coefficient | P value | β coefficient | P value | β coefficient | P value | β coefficient | P value |
| AD | Age | - | - | - | - | -0.12 | 0.13 | -0.13 | 0.10 |
|  | Sex | - | - | - | - | -0.12 | 0.16 | -0.08 | 0.31 |
|  | Education, years | -0.03 | 0.75 | -0.06 | 0.54 | 0.24 | 0.00 | 0.23 | 0.01 |
|  | Interscan interval | 0.23 | 0.03 | -0.08 | 0.43 | 0.40 | 0.00 | 0.40 | 0.00 |
|  | Baseline MMSE | -0.10 | 0.31 | 0.09 | 0.39 | -0.30 | 0.00 | - | - |
|  | Baseline DRS | - | - | - | - | - | - | -0.31 | 0.00 |
|  | vCSF growth | -0.09 | 0.41 | 0.32 | 0.00 | - | - | - | - |
| NC | Age | - | - | - | - | 0.35 | 0.02 | 0.38 | 0.01 |
|  | Sex | - | - | - | - | -0.33 | 0.01 | -0.37 | 0.01 |
|  | Education, years | -0.15 | 0.33 | -0.37 | 0.01 | -0.04 | 0.76 | -0.08 | 0.55 |
|  | Interscan interval | -0.13 | 0.37 | -0.03 | 0.83 | -0.10 | 0.45 | -0.08 | 0.55 |
|  | Baseline MMSE | -0.02 | 0.89 | 0.00 | 0.98 | -0.05 | 0.75 | - | - |
|  | Baseline DRS | - | - | - | - | - | - | 0.17 | 0.18 |
|  | vCSF growth | 0.35 | 0.02 | 0.48 | 0.00 | - | - | - | - |

Model 2: Similar findings were shown when assessing the vCSF growth with baseline DRS (β = -0.31, p < 0.001, **Figure 2b**), ISI (β = 0.40, p < 0.0001) and YOE (β = 0.23, p = 0.01). NCs also showed similar associations in age (β = 0.38, p = 0.01) and sex (β = -0.37, p = 0.01), but not in DRS (**Table 3**).

## Discussion

### Main finding

Our study found that AD displayed 14.6% annual ventricular growth as opposed to NCs who had only 11.8% annual growth. These numbers appear higher than those reported in the literature [1,23,24]; however, the present study used a newer imaging methodology to assess voxel-based progression as opposed to absolute volumetric difference (**Table 4**). Another possible explanation for these differences is that much of the recent literature has been based on the Alzheimer’s Disease Neuroimaging Initiative (ADNI), which has been considered to be a more “pure” AD sample with little vascular pathology in contrast to the “real-world” sample from the SDS [11].

**Table 4.**
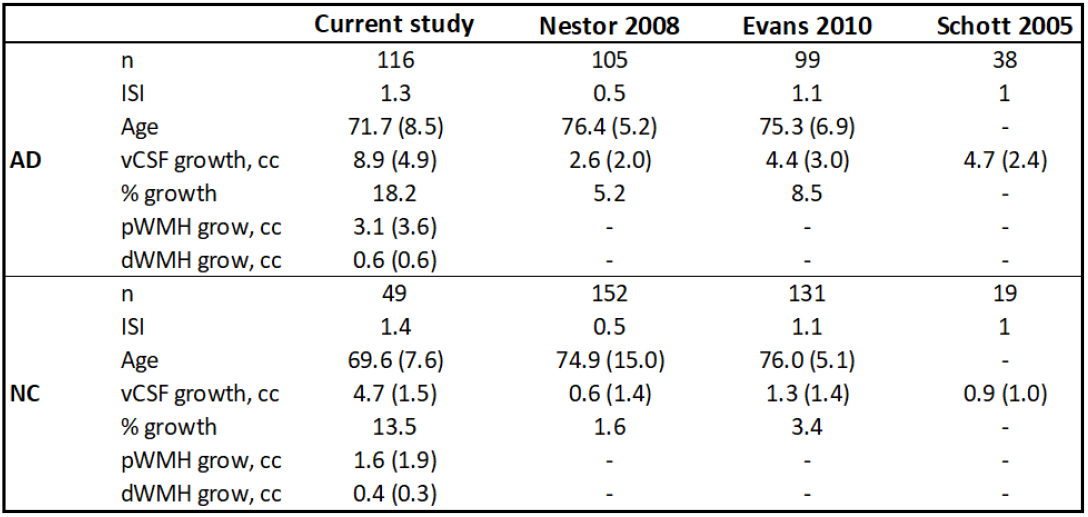
Present ventricular growth results compared with findings from previous research.

Interestingly, the AD showed a trend toward greater pWMH growth than the NCs while there was no difference in dWMH suggesting that these two pathologies may progress differently in different populations. The literature surrounding WMHs in AD typically uses global volumes rather than subdivided into dWMH and pWMH, making comparisons to the presently observed trend difficult. However, in cognitively healthy elderly adults, pWMH volume has been correlated with more reduced cerebral blood flow than dWMH [25]. Differences in dWMH and pWMH in AD should be the target of future research, preferably with a large sample size to increase power.

### Is vCSF growth associated with SVD burden?

The findings from the current study are consistent with previous research examining SVD and vCSF in AD. It has been shown that high SVD burden in AD is associated with larger ventricles [15] and greater vCSF growth over time [17] compared with NCs. A novel finding in the present study is that this relationship is not consistent with all types of SVD burden. In AD patients, greater vCSF growth was associated with greater pWMH while there was no significant difference in dWMH. To examine if severe periventricular white matter disease was driving these findings, we removed participants who had baseline pWMH volumes > 10cc (**Table 5**). The effect size for vCSF growth remained stable, in comparison to our entire sample (**Table 2**), providing support that the periventricular WMH load was not the main driving force for ventricular expansion. Additionally, an effect size calculation was calculated to determine the minimum number of participants required per group to detect a 25% change with alpha set to 0.05 at 80% power. It was determined that a minimum of 55 participants would be needed per group to detect this difference, although we would recommend 75 participants per group to account for 27% attrition.

**Table 5:** Demographic and volumetric data for participants with pWMH volumes < 10cc.findings from previous research.

|  | AD<br>(n = 88) | NC<br>(n = 42) | Cohen's d<br>effect size |
| --- | --- | --- | --- |
| **Baseline Demographics** |  |  |  |
| Age, years | 70 (9) | 68 (7) | 0.25 |
| Sex, n (M / F) | 40 / 48 | 15 / 27 |  |
| Insterscan Interval, years | 1.3 (0.4) | 1.4 (0.4) | 0.25 |
| MMSE/30<sup>a</sup> | 22.9 (2.9) | 28.9 (1.3) | 2.67 |
| DRS/144<sup>b</sup> | 118 (9.6) | 141 (2) | 3.32 |
| **Baseline Volumetrics** |  |  |  |
| BPF, % | 73.2 (4.4) | 79.2 (4.1) | 1.41 |
| vCSF, cc | 47 (20.1) | 32.3 (17) | 0.79 |
| pWMH, cc | 2.9 (2.8) | 1.6 (1.7) | 0.56 |
| dWMH, cc | 0.6 (0.8) | 0.5 (0.5) | 0.15 |
| **Growth Volumes** |  |  |  |
| vCSF, cc | 9.1 (5.3) | 4.6 (1.6) | 1.15 |
| pWMH, cc | 1.8 (2) | 1.1 (1.3) | 0.42 |
| dWMH, cc | 0.4 (0.4) | 0.3 (0.3) | 0.28 |

Although the relationship between the vCSF and SVD is not completely understood, there are a few proposed theories that attempt to explain their association. In a pathology study, pWMH but not dWMH were significantly associated with breakdown of the ventricular lining, arteriosclerosis, and neurofibrillary tangles [26], suggesting that location of WMHs may have different underlying causes. It has also been shown that there is collagenosis of the deep medullary veins surrounding the ventricles in AD leading to periventricular infarcts and edema [27]. We predict that over time, these infarcts begin to fuse into the vCSF increasing the growth rate relative to NCs. Additionally, it has been shown that the ventricles have a role in waste clearance in the brain and that AD patients have reduced clearance relative to controls [14]. The increased venous collagenosis in AD patients may also play a role in slowing down the vCSF filtration and ultimately in the rapid volumetric growth.

The link between vCSF growth and SVD can also be inferred from animal models and drug trials. AD has been linked with vascular risk factors such as hypertension [28] which correlates with greater cognitive impairment and neuropsychiatric symptoms [29]. In mouse models of AD, treating hypertension by use of an angiotensin receptor blocker (ARB) has been shown to improve cognitive functioning, reduce the inflammatory activation of astroglial and microglial cells, and some neurogenesis (although not statistically significance) [30]. This has led researchers to question whether treatments for hypertension, such as the use of an ARB, may slow ventricular atrophy (ClinicalTrials.gov ID: NCT02085265) and preserve cognitive functioning in human trials [29,31].

### Do baseline cognitive test scores predict ventricular growth?

It is well established that brain atrophy measured by ventricular growth is correlated with cognitive decline [6,32] especially in AD [1,23,33]. Additionally, brain atrophy has been shown to predict future cognitive decline and disease progression from normal aging to mild cognitive impairment to AD [34,35]. The present study has taken a novel look at this relationship, showing that baseline cognitive test scores predict more rapid ventricular atrophy and disease progression within AD. Both the baseline MMSE and DRS scores were strongly predictive of future vCSF enlargement, which supports the clinical utility of those tools. The MMSE and DRS are both used frequently in clinical settings to assess current cognitive functionality. With the addition of this study, clinicians can also use these scores to predict brain atrophy rates if no intervention is introduced.

## Conclusions

Our study findings demonstrate that real-world AD patients from a community-based memory clinic have significant and rapid ventricular expansion (∼15%/yr) that can be predicted using commonly used global cognitive tests (MMSE and DRS). This supports findings from other studies that recommend the use of vCSF expansion as an MRI-based AD biomarker of neurodegeneration in clinical trials. Additionally, we observed that in AD, ventricular expansion is more closely related to pWMH than dWMH, suggesting that periventricular vasculopathy plays a significant role in AD-related atrophy. As periventricular and deep WMH volumes are often combined as a single measure of vascular burden, future studies examining AD neurodegeneration should consider them as separate entities with different etiologies.

## Data Availability

The datasets generated and/or analysed during the current study are not publicly available due to consent-related issues conducting research embedded in care, however, data can be made available from the corresponding author upon request.

## Abbreviations

AD: Alzheimer’s Disease;
ADNI: Alzheimer’s Disease Neuroimaging Initiative;
ANOVA: analysis of variance;
BPF: brain parenchymal fraction;
(v/s) CSF: ventricular/sulcal cerebrospinal fluid);
DRS: Dementia Rating Scale;
ISI: interscan interval;
MMSE: Mini-Mental State Examination;
MRI: magnetic resonance imaging;
NAGM: normal appearing grey matter;
NAWM: normal appearing white matter;
NC: cognitively normal control;
NIA-AA: National-Institute on Aging-Alzheimer’s Association;
SDS: Sunnybrook Dementia Study;
ST-TIV: supratentorial-total intracranial volume;
SVD: cerebral small vessel disease;
(d/p) WMH: deep/periventricular white matter hyperintensity;
YOE: years of education

## Declarations

### Ethics approval and consent to participate

This research was ethically conducted on consenting study participants and approved by the Sunnybrook Research Ethics Board.

### Consent for publication

Not applicable

## Authors’ contributions

SA: Conceptualisation, Data Curation, Formal Analysis, Investigation, Methodology, Project Administration, Validation, Visualisation, and Writing (draft, review, and editing)

JR: Conceptualisation, Data Curation, Formal Analysis, Investigation, Methodology, Software, Validation, Writing (draft, review, and editing), and Supervision

MFH: Data Curation, Formal Analysis, Writing (draft, review, editing)

FG: Data Curation, Formal Analysis, Writing (review, editing), Supervision

LZ: Project Administration, Writing (review), Resources

MM: Conceptualisation, Supervision, Writing (review and editing), Resources, Funding Acquisition

SEB: Conceptualisation, Methodology, Supervision, Writing (review and editing), Resources, Funding Acquisition

## Acknowledgements

We would like to thank the participants who have graciously donated their time, consent, and participation in the Sunnybrook Dementia Study. Thank you to the Linda C. Campbell foundation, the analysts, database experts, software developers, the BrainLab.ca neuroimaging team, psychometrists, and the cognitive neurology clinical team for their ongoing herculean efforts and contribution to this study.

